# Workplace ventilation improvement to address coronavirus disease 2019 cluster occurrence in a manufacturing factory

**DOI:** 10.1101/2022.04.04.22271935

**Authors:** Hiroko Kitamura, Yo Ishigaki, Hideaki Ohashi, Shinji Yokogawa

**Affiliations:** Occupational Health Training Center, University of Occupational and Environmental Health, Japan, Kitakyushu, Fukuoka, Japan; Graduate School of Informatics and Engineering, University of Electro-communications, Chofu, Tokyo, Japan; Info-powered Energy system Research Center (i-PERC), University of Electro-communications, Chofu, Tokyo, Japan

## Abstract

**Aim and Methods:** A coronavirus disease 2019 (COVID-19) cluster emerged in a manufacturing factory in early August 2021. In November 2021, a ventilation survey using tracer gas and data analysis was performed to reproduce the situation at the time of cluster emergence and verify that ventilation in the office increased the risk of aerosol transmission; verify the effectiveness of measures implemented immediately in August; and verify the effectiveness of additional measures when previously enforced measures proved inadequate.

**Results:** At the time of cluster emergence, the average ventilation frequency was 0.73 times/h, less than the 2 times/h recommended by the Ministry of Health, Labour, and Welfare; as such, the factory’s situation was deemed to have increased the risk of aerosol transmission. Due to the measures already taken at the time of the survey, the ventilation frequency increased to 3.41 times/h on average. It was confirmed that ventilation frequency increased to 8.33 times/h on average, when additional measures were taken.

**Conclusion:** To prevent the re-emergence of COVID-19 clusters, it is necessary to continue the measures that have already been implemented. Additionally, introduction of real-time monitoring that visualizes CO_2_ concentrations is recommended. Furthermore, we believe it is helpful that external researchers in multiple fields and internal personnel in charge of health and safety department and occupational health work together to confirm the effectiveness of conducted measures, such as this case.

## Introduction

Since a case of unexplained pneumonia was reported in Wuhan City, China, in December 2019, coronavirus disease 2019 (COVID-19) has spread across the globe. By mid-December 2021, at least 272,819,000 global COVID-19 cases were reported, and 5,636,000 people were confirmed dead as a result. On January 8, 2020, the United States Center for Disease Control (CDC) officially announced that the cause of lung inflammation related to the seafood market in Wuhan, China, was the SARS-CoV-2 virus.

Initially, infection with SARS-CoV-2 was caused by exposure to airway secretions containing the virus, and the transmission route was thought to be contact and droplet infection. In May 2021, based on the scientific knowledge accumulated to date, the CDC declared that the transmission routes of SARS-CoV-2 are as follows: (1) aerosol transmission (inhalation of air containing fine droplets and aerosol particles), (2) droplet transmission (adherence of droplets and fine particles to the mucous membranes), and (3) contact transmission (contact of mucous membranes with fingers contaminated with airway secretions containing the virus or with those that had been in contact with surfaces contaminated with the virus). An outbreak of COVID-19 in poorly ventilation restaurant has been reported^1^. At the end of October 2021, the Japanese Ministry of Health, Labour, and Welfare (MHLW) stated that transmission occurs when fine particles (aerosol) in the air containing SARS-CoV-2 are inhaled; as such, the Ministry recommends ensuring proper ventilation to combat aerosol transmission.

Japan has “public health centers” which are public institutions under the jurisdiction of the MHLW. There are 469 such centers nationwide based on the Community Health Act. These public health centers provide a comprehensive professional and technical base that supports the health of residents by providing consultation on intractable diseases and mental health, implementing infection countermeasures, and conducting monitoring and guidance on pharmaceutical affairs, food hygiene, and environmental hygiene. They also form a base for health crisis management, such as preventing the spread of diseases in the event of a health crisis and disseminating relevant information. Public health centers conduct “active epidemiological investigation” on patients with confirmed COVID-19 detected in Japan following the Act on the Prevention of Infectious Diseases and Medical Care for Patients with Infectious Diseases. In these investigations, the patient is asked about their activities within 14 days prior to the onset of symptoms to estimate the source and route of infection. They are also asked about their activities 2 days prior to the onset of symptoms to identify close contacts.

In the office of a manufacturing factory in Fukuoka Prefecture, five confirmed cases of COVID-19 successively emerged within 1 week in early August 2021, and the infections were confirmed to be a cluster (a group of infected people whose infections are linked). The local public health center stated that “the infected people were concentrated inside the office, and although there are working electric fans inside, there are no vents through which the virus can escape, resulting in the spread of infection.” Within August, the office ventilation equipment was inspected and repaired to improve these issues, electric fans were relocated to the opposite side of the room considering the airflow, and the room door was opened when working inside. However, as mentioned above, the public health center investigation was based on interviews, and the office’s ventilation when the cluster emerged was not evaluated. The level of ventilation after improvement measures were taken was not evaluated either.

The objective of this study is to: (1 reproduce the environmental variables at the time of cluster emergence and verify that ventilation in the office increased the risk of aerosol transmission; (2) verify the effectiveness of measures taken in August (inspection and repair of ventilation equipment in the office, relocation of electric fans, opening of doors during work) in response to the emergence of the cluster; and (3) verify the effectiveness of additional measures (opening windows, opening doors at the end of corridors, opening doors in adjacent rooms) when previously enforced measures proved inadequate.

## Materials and Methods

### Workplace conditions where the cluster emerged

The affected workplace is a factory and performs desk work in the office and intervention on the operation site. It is managed by mixed groups of daytime workers and two groups with two shifts. On average, workers spend 70% of their time in the office and 30% in the field out of the 7 hours and 45 minutes of work per day. At the time of the cluster’s emergence, 2–3 people were working in the daytime, and 16 people were working in shifts. As a measure against droplet transmission, 1.4m high partitions had been installed between desks. Layout of the workplace was shown in Figure 1.

**Figure 1:** This figure is available at https://doi.org/10.6084/m9.figshare.19465091.v1.

### Status of cluster emergence

In early August, 2021, the first patient, indicated as P1 in Figure 2, was confirmed by polymerase chain reaction ^1^, and then PCR-positive individuals emerged in August within the same group (P2 to P5 in Figure 2 in order). Patients were found only in one of the two groups of shift workers. Interviews with workers revealed the following: P1, P2 and P3 had many opportunities to talk to each other in close proximity in the aisles between desks, P4 and P5 had many opportunities to be in close proximity to other patients not only in the office but also outside of the office, and P1 to P5 often worked in close proximity and talked while they all looked at the same monitor. Figure 3 showed the time course of the emergence of infected patients and physical condition of the patients.

**Figure 2,3:** This figure is available at https://doi.org/10.6084/m9.figshare.19465091.v1.

### Ventilation frequency survey method

The office’s ventilation frequency was measured as follows:

1. Eight CO_2_ sensors, indicated as S1 to S8 in Figure 4, were installed. One sensor was installed for each section, separated by the infected person’s desk and partition. The CO_2_ sensor measured and recorded the average CO_2_ concentration every 60 s. A non-dispersive infrared absorption-type CO_2_ sensor, TR-76Ui (T&D, Matsumoto, Japan), was used for the measurement. The TR-76Ui sensor can detect CO2 concentrations from 0 to 9,999 ppm, with an accuracy of ±50 ppm (±5%).
2. The office’s air conditioner and ventilation system were turned off, and all windows and doors were closed.
3. Dry ice was then vaporized to fill the office with CO_2_, raising the CO_2_ concentration well above the background atmospheric CO_2_ concentration (400 ppm).
4. Ventilation equipment, electric fans, windows, and doors were set under experimental condition1 shown in Table 1, and at the same time, all employees and researchers left the office because they were CO_2_ sources. The time at this point was considered as the ventilation start time.
5. The CO_2_ concentration’s transition was monitored remotely, and the measurement was completed after confirming that the CO_2_ concentration had dropped sufficiently.
6. For the next survey under experimental condition 2 shown in Table 1, the office’s air conditioner and ventilation system were turned off, and all windows and doors were closed.
7. Dry ice was then vaporized to fill the office with CO_2_, raising the CO_2_ concentration well above the background atmospheric CO_2_ concentration (400 ppm).
8. Ventilation equipment, electric fans, windows, and doors were set under experimental condition 2, and at the same time, all employees and researchers left the office because they were CO_2_ sources. The time at this point was considered as the ventilation start time.
9. The CO_2_ concentration’s transition was monitored remotely, and the measurement was completed after confirming that the CO_2_ concentration had dropped sufficiently.
10. For the next survey under experimental condition 3 shown in Table 1, the office’s air conditioner and ventilation system were turned off, and all windows and doors were closed.
11. Dry ice was then vaporized to fill the office with CO_2_, raising the CO_2_ concentration well above the background atmospheric CO_2_ concentration (400 ppm).
12. Ventilation equipment, electric fans, windows, and doors were set under experimental condition 3, and at the same time, all employees and researchers left the office because they were CO_2_ sources. The time at this point was considered as the ventilation start time.
13. The CO_2_ concentration’s transition was monitored remotely, and the measurement was completed after confirming that the CO_2_ concentration had dropped sufficiently. *Estimation of ventilation frequency*

**Figure 4:** This figure is available at https://doi.org/10.6084/m9.figshare.19465091.v1.

**Table 1.** Experimental conditions

| Factor<sup>a</sup> |  | Experimental<br>condition 1: during<br>cluster emergence | Experimental<br>condition 2: after<br>improvement<br>measures | Experimental<br>condition 3:<br>additional measures |
| --- | --- | --- | --- | --- |
| 1 | air conditioner | on | on | on |
| 2 | air conditioner | on | on | on |
| 3 | electric fan<sup>b</sup> | on | on | on |
| 4 | ventilation fan | off | on | on |
| 5 | ventilation fan | off | off | on |
| 6 | total heat<br>exchange type<br>ventilator | off | on | on |
| 7 | door | 30-mm open | fully open | 30-mm open |
| 8 | door | closed | closed | 30-mm open |
| 9 | door | closed | fully open | fully open |
| 10 | door | closed | closed | 30-mm open |
| 11 | window | closed | closed | 100-mm open |

a: Factor 1 to 11 in the Table 1 were linked to 1 to 11 in Figure 4.
b: Electric fans were placed in different locations in experimental condition 1 and experimental condition 2 and 3, as shown in Figure 4.

Based on the data measured by the CO_2_ sensor, the ventilation frequency was estimated using Seidel’s formula^2, 3^:

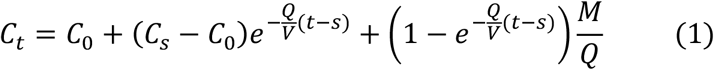

Where *C*: Indoor pollutant concentration (CO_2_ concentration in this study); *C*_0_: Steady-state value in the absence of pollution (400 ppm with atmospheric CO_2_ concentration as the background in this study); *V*: Room volume; *Q*: Ventilation volume; *M*: amount of pollutants generated; *t, s*: Time (Cs is the CO_2_ concentration at the ventilation start time in this study). When investigating the ventilation frequency, the room was unoccupied, and there was no other source of CO_2_. Therefore, if M = 0, Equation 1 can be transformed as follows:

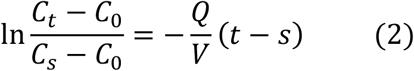

Based on the Equation 2, the ventilation frequency 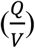 was estimated from the slope of the natural logarithm of the CO_2_ concentration decrease ratio with respect to the elapsed time from the ventilation initiation (ventilation time).

### Statistical analysis

We used the mixed-effect model to analyze the effects of experimental conditions on ventilation and the differences and trends in ventilation frequency related to CO_2_ sensor location. The dependent valuable was the ratio of the increase in CO_2_ concentration from the background to that at the ventilation initiation. We treated the ventilation time and interaction of ventilation time and the experimental conditions and interaction of ventilation time and CO_2_ sensor location as fixed effects, and the experimental condition and the sensor location as random effects. We used the statistical software JMP Pro Ver. 16 (JMP Statistical Discovery LLC.).

### Ethical consideration

This study was approved (approval number 21005) by the Ethics Committee on Experiments on Human Subjects of the University of Electro-Communications, Chofu, Tokyo, Japan.

## Results

Table 2 shows the estimated ventilation frequency calculated for each sensor under each experimental condition. The mean estimated ventilation frequency of sensors 1–8 was 0.74 times/h when the cluster emerged: experimental condition 1, 3.41 times/h after improvement measures in experimental condition 2, and 8.33 times/h for additional measures in experimental condition 3.

**Table 2.** Estimated ventilation frequency per sensor

| Sensor | Estimated ventilation frequency ± 95% CI<sup>a</sup> (times/h) | | |
| --- | --- | --- | --- |
|  | Experimental condition 1 | Experimental condition 2 | Experimental condition 3 |
| S1 | $0.726 \pm 0.008$ | $3.307 \pm 0.033$ | $8.673 \pm 0.261$ |
| S2 | $0.728 \pm 0.007$ | $3.332 \pm 0.040$ | $8.479 \pm 0.202$ |
| S3 | $0.721 \pm 0.007$ | $3.316 \pm 0.034$ | $8.332 \pm 0.191$ |
| S4 | $0.726 \pm 0.008$ | $3.292 \pm 0.031$ | $8.509 \pm 0.222$ |
| S5 | $0.754 \pm 0.008$ | $3.431 \pm 0.080$ | $8.598 \pm 0.174$ |
| S6 | $0.761 \pm 0.007$ | $3.578 \pm 0.043$ | $8.458 \pm 0.169$ |
| S7 | $0.726 \pm 0.008$ | $3.442 \pm 0.059$ | $7.666 \pm 0.123$ |
| S8 | $0.759 \pm 0.012$ | $3.580 \pm 0.062$ | $7.932 \pm 0.158$ |
| mean | 0.738 | 3.410 | 8.331 |

a: CI: confidence interval

The effects of the ventilation time on the CO_2_ concentration and the interaction of the ventilation time and experimental conditions were statistically significant (p<0.01). In other words, the CO_2_ concentration was significantly reduced by greater ventilation, and the ventilation frequency was significantly different depending on the experimental conditions (Table 3). On the other hand, the effect of the interaction of the ventilation time and the CO_2_ sensor location was not statistically significant (p=0.35), and there was no obvious difference in the ventilation frequency depending on the sensor location. Since the individual effects of the experimental conditions and the CO_2_ sensor location, which are random effects, were not statistically significant (p>0.05), respectively, it was unlikely that there was an individual effect other than the effects on the ventilation frequency (Table 3).

**Table 3.** Effects of experimental conditions and CO_2_ sensor location by mixed-effect model

| Factor | Number of parameters | DoF<sup>a</sup> of numerator | DoF<sup>a</sup> of denominator | F-value | p-value |
| --- | --- | --- | --- | --- | --- |
| Ventilation time<sup>b</sup> | 1 | 1 | 716.0 | 43036.35 | <0.01 |
| Interaction of Ventilation time and Experimental conditions | 2 | 2 | 716.0 | 12797.13 | <0.01 |
| Interaction of Ventilation time and CO<sub>2</sub> sensor location | 7 | 7 | 716.0 | 1.13 | 0.35 |

| Dispersion component | Estimated value | Standard error | 95% CI<sup>a</sup><br>Lower limit | 95% CI<sup>a</sup><br>Upper limit | Wald p-value |
| --- | --- | --- | --- | --- | --- |
| Experimental conditions | 1.042 | 1.042 | −1.000 | 3.085 | 0.317 |
| CO<sub>2</sub> sensor location | 0.000296 | 0.000177 | −0.000051 | 0.000642 | 0.0945 |
| Residual error | 0.00323 | 0.000171 | 0.00292 | 0.00359 |  |
| sum | 1.046 | 1.042 | 0.284 | 40.398 |  |

a: CI: confidence interval

## Discussion

According to experimental condition 1, the ventilation frequency at the time of cluster emergence was 0.73 times/h on average. Menzies et al.^4^ reported that a lack of ventilation is associated with an increased incidence of airborne infections; the higher the ventilation frequency, the higher the efficiency of air dilution, and the lower the risk of airborne infection. They also stated that a ventilation frequency of fewer than 2 times/h is associated with the spread of tuberculosis, an airborne infection. It has also been reported that the ventilation frequency in junior high school classrooms where tuberculosis outbreaks occurred in Japan ranger between 1.6–1.8 times/h.^5^ According to the Guide for Outpatient Treatment of COVID-19 by the Japan Medical Association, aerosols containing the SARS-CoV-2 virus from infected people will remain airborne for more than 3 h in an unventilated room.^6^ Strictly speaking, there is a difference between airborne transmission and aerosol transmission; however, a ventilation frequency of only 0.73 times/h identified in this study increased the risk of aerosol transmission. The mean ventilation frequency in experimental condition 2 was 3.41 times/h, 4.7 times higher than when the cluster emerged. Under experimental condition 3, the mean ventilation frequency was 8.33 times/h, 11.4 times higher than when the cluster occurred. The MHLW stated that to improve the ventilation of closed rooms with poor ventilation, the ventilation frequency should be increased to ≥2 times/h by opening windows without using mechanical ventilation (e.g., air conditioning and mechanical ventilation equipment).^7^ Under experimental condition 2, it was confirmed that the minimum ventilation frequency was secured.

The standard CO_2_ concentration according to the Act on Maintenance of Sanitation in Buildings is ≤1,000 ppm, and it is necessary to secure an outside air introduction amount of approximately ≥30 m^3^/h per person to manage a space’s CO_2_ concentration at ≤1,000 ppm. Since the volume of the office examined in this study was about 240 m^3^, in order to manage CO_2_ concentration in the office at ≤1000 ppm or less at the average ventilation frequency of 0.73 and 3.41 times/h in experimental conditions 1 and experimental condition 2, respectively, it was estimated that the ventilation achieved under experimental condition 1 could cover five people while that of experimental condition 2 can cover 27 people.

At the time of the cluster emergence, 18–19 people, including daytime workers and shift workers, were working simultaneously. The ventilation volume was <30% of the required amount, which increased the risk of aerosol transmission. The ventilation volume achieved under experimental condition 2 would allow the original number of employees to work inside the office, and that under experimental condition 3 would allow 66 people to work in the office simultaneously. Of course, the amount of CO_2_ that humans exhale depends on their level of physical activity.^8^ As such, it would be safer to accommodate a smaller number of people than the above estimation when heated discussions takes places or when there is heavy activity to simulate the operation of production line. If the door cannot be released to discuss sensitive content, it also would be safer to keep the number of people inside small and the meeting time as short as possible.

In our previous investigation of a cluster occurrence,^9^ we reported that 1.6-m high vinyl sheet partitions installed between desks facing each other as a measure against droplet transmission blocked the office’s airflow, resulting in a section where air stagnated (vinyl sheet cluster). In the office space of this study, there were no differences in ventilation frequency depending on the sensor location, and there seemed no inhibitory effects of inappropriate partitions on ventilation. However, in the office, an electric fan created an airflow, as shown from the lower side to the upper right part of Figure 1, which considered to help scatter droplets containing the virus released from the first infected person without an outlet for air to escape to the outside. As a result, it was suggested that virus-containing droplets had gradually accumulated in the upper right part of the office shown in Figure 1. The office’s ventilation was extremely poor at <1 time/h, making it possible for a “leeward cluster” to emerge. In order to prevent leeward clusters, when using an electric fan or blower to ensure adequate air circulation in a room, it is necessary to secure an air outlet and create outflow.

The following two proposals may be feasible for the operation of the office: First, when doing regular work, set experimental condition 2. If more people are required to work inside the office or during long discussions, add the countermeasure plan confirmed in experimental condition 3. Second, maintain experimental condition 2 and limit the number of people entering the room according to the work content. The use of CO_2_ sensors to control indoor air quality has attracted significant attention. In addition to the measures mentioned above, introduction of real-time monitoring that visualizes CO_2_ concentrations, which can be used to determine the timing of ventilation and limit the number of people entering the room, is recommended.^10-12^ We believe that CO_2_ concentration visualization can create an environment that is more flexible and allows workers to work with greater peace of mind.

It is essential to improve ventilation in the workplace, considering feasibility and sustainability and measures that can be put into practice without impeding work. The measures proposed in this study are based on workplace improvement activities and could be implemented continuously without difficulty. Of course, there are cases where improvements led by experts and researchers are necessary, but in the long run, the measures taken by workers who use the site daily are considered essential to prevent the recurrence of COVID-19 clusters.

## Conclusion

The survey of ventilation of the manufacturing factory where a COVID-19 disease cluster emerged revealed that: (1) the average ventilation frequency in the office at the time of cluster emergence was 0.73 times/h, which increased the risk of aerosol transmission; (2) the improvement measures already implemented in the office, which increased the average ventilation frequency to 3.41 times/h were effective; and (3) the average ventilation frequency after implementing additional measures was 8.33 times/h, which was 2.4 times that after the improvement measures, suggesting that ventilation could be further improved. In the future, the introduction of real-time monitoring that visualizes CO_2_ concentrations while improving ventilation and controlling the number of occupants is recommended. In order to prevent the spread of novel coronavirus infection in the workplace, employers would take measures considered to be effective. Additionally, it is helpful that external researchers in multiple fields and internal personnel in charge of health and safety department and occupational health work together to confirm the effectiveness of conducted measures, such as this case. Ventilation is just one measure against aerosol transmission. As measures against COVID-19 in the workplace, it is necessary to continue to avoid the three Cs (closed spaces, crowded places, and close-contact settings), ensure thorough hand hygiene, universal mask-wearing, and social distancing.

## Data Availability

All data produced in the present study are available upon reasonable request to the authors.

## Acknowledgement

This work was supported by JSPS KAKENHI Grant No. 21K19820.

HK: Methodology, Investigation, Writing-Original draft preparation, YI: Methodology, Investigation, Writing-Reviewing and Editing, HO: Investigation, SY: Methodology, Analysis, Writing-Reviewing and Editing.

This study was approved (approval number 21005) by the Ethics Committee on Experiments on Human Subjects of the University of Electro-Communications, Chofu, Tokyo, Japan. The authors declare no conflicts of interest associated with this manuscript. This work was supported by a KAKENHI Grant (No. 21K19820) from JSPS.

